# Serum SARS-CoV-2 Antigens for the Determination of COVID-19 Severity

**DOI:** 10.1101/2021.11.18.21266478

**Authors:** Julien Favresse, Jean-Louis Bayart, Clara David, Constant Gillot, Grégoire Wieërs, Gatien Roussel, Guillaume Sondag, Marc Elsen, Christine Eucher, Jean-Michel Dogné, Jonathan Douxfils

**Author notes:** **Correspondence:** Jonathan Douxfils, University of Namur, Namur, Belgium, B-5000 Namur - Belgium.

## Abstract

The diagnostic of SARS-CoV-2 infection relies on reverse transcriptase polymerase chain reactions (RT-PCR) performed on nasopharyngeal (NP) swabs. Nevertheless, false negative results can be obtained with inadequate sampling procedures making the use of other matrices of interest. This study aims at evaluating the kinetic of serum N antigen in severe and non-severe patients and compare the clinical performance of serum antigenic assays with NP RT-PCR. Ninety patients were included and monitored for several days. Disease severity was determined according to the WHO clinical progression scale. The serum N antigen was measured with a chemiluminescent assay (CLIA) and the Single Molecular Array (Simoa). Thresholds for severity were determined. In severe patients, the peak antigen response was observed 7 days after the onset of symptoms followed by a decline. No peak response was observed in non-severe patients. Severity threshold for the Simoa and the CLIA provided positive likelihood ratio of 30.0 and 10.9 for the timeframe between day 2 and day 14, respectively. Compared to NP RT-PCR, antigenic assays were able to discriminate the severity of the disease (p = 0.0174, 0.0310 and p = 0.1551 with the Simoa, the CLIA and the NP RT-PCR, respectively). Sensitive N antigen detection in serum thus provides a valuable new marker for COVID-19 diagnosis and evaluation of disease severity. When assessed during the first 2 weeks since the onset of symptoms, it may help in identifying patients at risk of developing severe COVID-19 to optimize better intensive care utilization.

## Introduction

The diagnostic of SARS-CoV-2 infection still relies on molecular assays with reverse transcriptase polymerase chain reaction (RT-PCR) performed on nasopharyngeal (NP) swabs being considered as the gold standard detection method.(1) Other techniques like chemiluminescent immunoassays (CLIA) or Single Molecular Array (Simoa) have demonstrated good correlation with RT-PCR on NP swabs at least for cycle thresholds (Ct) below 33.(2, 3) Other matrices like saliva, plasma or dried blood spots have been used in order to detect SARS-CoV-2 infection and also revealed interesting performance versus RT-PCR.(4-6) Nevertheless, the clinical performance depends on the detection methods. Namely, lateral flow assay (LFA) permits to provide results within a couple of minutes, but their performance is relatively weak with some rapid antigen detection (RAD) assays showing clinical sensitivity below 30%.(7) However, while almost all RT-PCR techniques show excellent sensitivity with various methods demonstrating limit of detection (LOD) from 10^2^ to 10^5^ copies/mL according to manufacturer package inserts and reference panels,(8) the correlation between RT-PCR results from NP swabs and the disease severity has been questioned.(9, 10) The Ct values for a specimen vary between different kits and techniques (including target genes, primers and threshold fluorescence values) and Ct values may vary between different runs of the same kit.(10) The Ct value also depends on the method of collection of the sample and hence there may be a variation in Ct values between two different samples obtained from the same person on the same day and run on the same kit.(11) In addition, concerns have been raised about the risk of false-negative results associated with the use of nasal and NP swabs, especially before symptom onset.(12) Thus, there is a place for improvement in the diagnostics of SARS-CoV-2 infection, the “ideal” test being able to detect the presence of the virus with a similar or better sensitivity than the RT-PCR performed on NP swabs, but with a better prognostic value regarding the severity of the disease with a more convenient access to biological specimen.(13, 14)

Blood samples are used to determine the antibody production and the adaptative immunity but are usually less used for the diagnosis of acute infection, especially in respiratory disease because of the focal nature of the infection or possible pre-existence of antibodies.(15) Nevertheless, antigen detection in non-respiratory fluids is still used for two respiratory bacteria: Streptococcus pneumonia and Legionella pneumophila. Interestingly, it has also been reported in SARS-CoV-1 infection.(16) Furthermore, multiple clinical manifestations suggest that SARS-CoV-2 can migrate from the lungs into the bloodstream.(5, 17) This study aims at evaluating the clinical performance of two serum antigen assays for the diagnostic of SARS-CoV-2 infection and prognostic determination of the disease severity.

## Material and methods

### Study Design and Patients Selection

All patients with documented molecular diagnosis of SARS-CoV-2 infection between April 2020 and July 2021 with at least one blood sample collected within one day of the NP collection were potentially includible. One patient may have multiple blood collection after the diagnosis and SARS-CoV-2. Minimal medical information required days since the onset of symptoms and sufficient information to establish the evaluation of the disease severity according to the WHO clinical progression scale.(18) Reviewers of the medical records and technicians who performed the analyses were blinded and not allow to communicate on the results. Based on these criteria, a total of 92 SARS-CoV-2 infected patients were retrospectively retrieved from our laboratory biobank, representing 250 serum samples. Medical records were sufficiently documented for 90 patients. None of the patients were vaccinated against SARS-CoV-2. The 2 patients with insufficient clinical information (i.e. no information regarding days since symptom onset or clinical outcomes) were not included for analyses. The remaining 90 patients, for whom 243 samples were available, represented the study population. Serum samples of these patients were analyzed for evaluation of the antigen kinetics. Among these 90 patients, 84 had blood samples collected between days 2 and 14 since the onset of symptoms. These 84 patients were included for the determination of a cut-off for the determination of disease severity and represented the subpopulation A. For 81 out of the 90 patients, a blood sample was obtained within 12 hours of the NP sampling allowing the evaluation of the performance of serum antigen versus NP RT-PCR detections in similar temporal condition from infection onset. These 81 patients represented the subpopulation B (**Supplemental Figure 1**). Note that patients may be included in both subpopulation A and B if they responded to the criteria of the respective subpopulations. Out of the 90 patients, 11 patients were categorized as asymptomatic (WHO score of 1), 17 as symptomatic ambulatory mild disease (WHO score of 2 and 3), 47 as symptomatic hospitalized moderate disease (WHO scores of 4 and 5) and 15 as symptomatic hospitalized severe diseased (WHO score of 6 to 10). Twelve patients died (WHO score of 10). Description of the cohort and inclusion of the patients and corresponding samples in the different part of this study is reported in **Figure 1**. Demographic data of the population is presented in **Supplemental Table 1**. Among the 90 included patients, 44 (48.9%) were females (median age = 78 years, min-max = 20-97) and 46 (51.1%) were male (median age = 77 years, min-max = 33-94). A cohort of 71 pre-pandemic serum samples collected before February 2020 was also included to evaluate the specificity of antigen assays. The samples were retrieved from our laboratory biobank. The study protocol is available upon request and was in accordance with the Declaration of Helsinki and approved by the Medical Ethical Committee of Saint-Luc Bouge (Bouge, Belgium, approval number B0392020000005).

**Figure 1:**
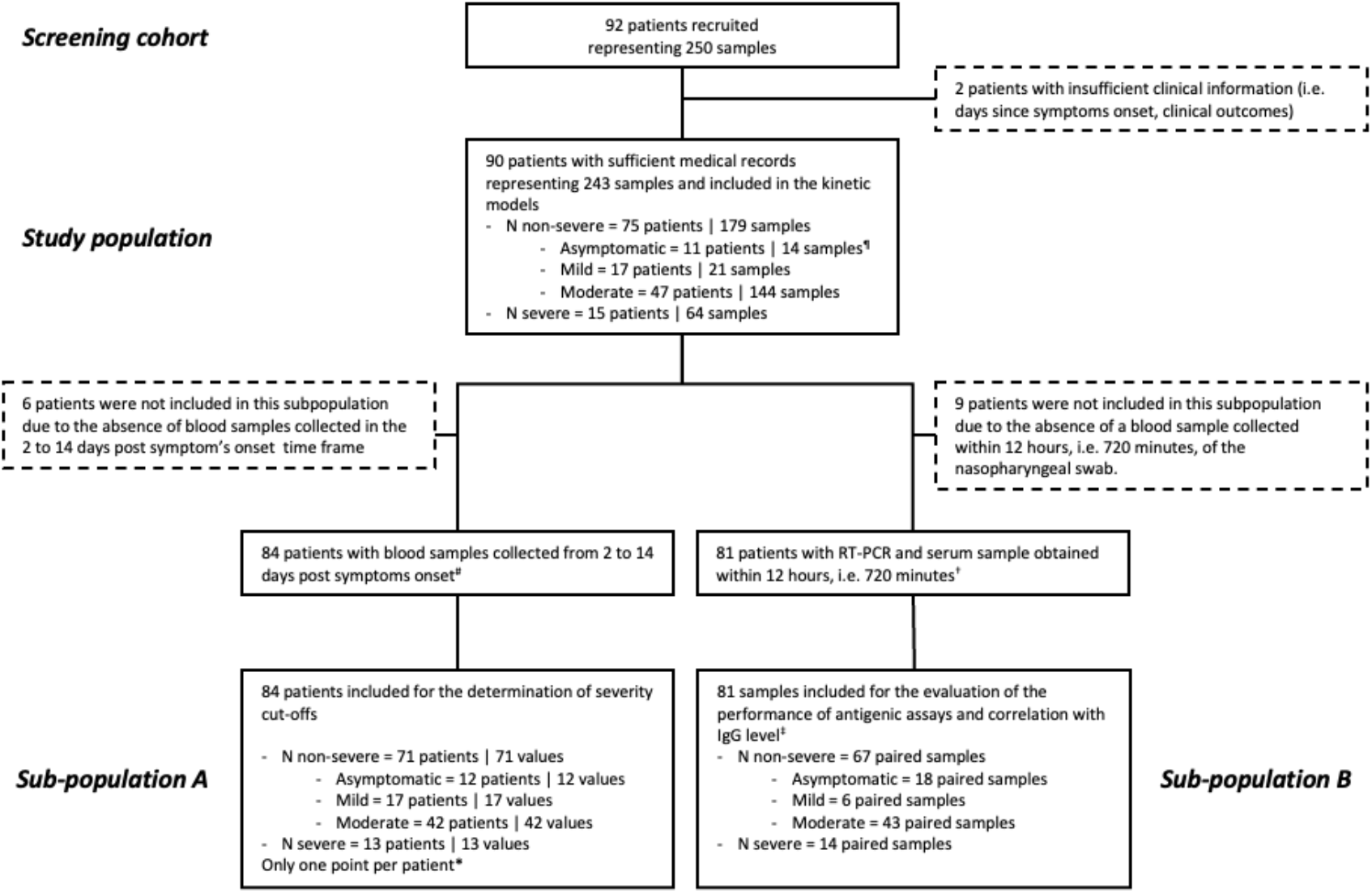
Study flow diagram. † Blood samples and RT-PCR swabs of 9 patients were more than 720 minutes apart. Ninety percent of the samples were obtained within 3 hours. ‡ SARS-CoV-2 Spike IgG measurement was only performed in 77 out of the 81 samples due to insufficient biological material. Immunoglobulin G measurement was only performed on those 77 samples. ¶ In some kinetic models, asymptomatic patients and patients with positive SARS-CoV-2 spike immunoglobulin G were excluded. * Multiple sample inclusion criteria were applied to investigate different possible cut-off for severity: a) earliest antigenemia value since symptom onset within the day 2 to day 14 window for a particular patient, b) mean of all antigenemia values of samples collected within the day 2 to day 14 window for a particular patient and c) the maximal antigenemia value obtained within the day 2 to day 14 window. All these inclusion strategies reported one value per patient. # Six patients had blood samples collected beyond 14 days since symptom onset.

### Nasopharyngeal and Blood Collection

Nasopharyngeal samples were collected using eSwab liquid preservation medium tubes (Copan Italia, Brescia, Italy) and analyzed by RT-PCR without any delay. Blood samples were collected in serum-gel tubes (BD SST II Advance®, Becton Dickinson, NJ, USA) and centrifuged for 10 min at 1740×g on a Sigma 3-16KL centrifuge. Sera were stored in the laboratory serum biobank at −20°C from the collection date. Frozen samples were thawed for 1 hour at room temperature on the day of the antigen analysis. Re-thawed samples were vortexed before the analysis.

### Antigen Assays

#### Single Molecular Array

The SARS-CoV-2 nucleocapsid (N) antigen was quantified automatically in patient sera by a Single Molecular Array (Simoa) immunoassay using the Simoa HD-X analyzer (Quanterix, Massachusetts, USA). Samples were analyzed using the commercial SARS-CoV-2 N-Protein Advantage kit (item 103806), a paramagnetic microbead-based sandwich ELISA. Briefly, diluted sample and anti-N protein antibody coated capture beads and detector antibodies are combined for 35 minutes in the cuvette in a first step. Beads are then washed, and a conjugate of streptavidin-ß-galactosidase is added to label the captured N protein. After washing, beads are resuspended with resorufin-ß-galactopyranozide for signal generation. Finally, beads are loaded in microarrays of femtoliter reaction wells. The fraction of bead-containing microwells counted with an enzyme is converted into “average enzymes per bead” (AEB). AEB values are converted into N protein concentration by interpolation from the calibration curve obtained by 4-parameter logistical regression fitting. The results are quantitative and expressed as pg/mL. This assay uses 8 calibrators ranging from 0 pg/mL to 200 pg/mL. Serum samples that provided results upper the calibration range were retested with a 1,000-fold dilution. The LOD of the assay is 0.099 pg/mL. The within-run coefficient of variation (CV) is less than 10%. Positivity cut-offs in serum samples were not disclosed by the manufacturer.

#### iFlash-2019-nCoV Assay

The SARS-CoV-2 N antigen was detected automatically in patient sera by CLIA using the iFlash 1800 automated magnetic CLIA (MCLIA) analyzer (Shenzhen YHLO Biotech Co. Ltd., Shenzhen, China). Samples were analyzed using the commercial iFlash-2019-nCOV Antigen assay kit. Antigens in the sample will react with anti-2019-nCoV antibodies coated on paramagnetic particles and with anti-2019-nCoV acridinium-ester-labeled conjugate to form a sandwich complex. Under magnetic field, particles are absorbed to the wall of the reaction chamber, and unbound materials are washed away. Afterwards, the Pre-Trigger and Trigger are added to the reaction mixture. The chemiluminescent signal is then measured as relative light units (RLUs). Results are determined via a 2-point calibration curve. The results are only qualitative and are expressed as a cut-off index (COI). The within-run CV ranges from 2.7 to 3.6%.(19) The manufacturer’s positivity cut-off is > 1.0 COI. The indented use only includes the detection of antigens in NP swabs. In our study, we explored the possibility of detecting the antigen in serum (RUO setting).

### SARS-CoV-2 Spike IgG

SARS-CoV-2 Spike IgG antibodies were quantified automatically in patient sera by a Simoa immunoassay using the Simoa HD-X analyzer (Quanterix, Massachusetts, USA). Samples were analyzed using the commercial SARS-CoV-2 Spike IgG Advantage kit (item 103769), using a similar procedure as for the SARS-CoV-2 N antigen. The positivity cut-off of 924 ng/mL corresponding to the maximal value obtained in pre-pandemic serum samples was used.(20) SARS-CoV-2 antibodies were available for 77 patients out of the 81 (95.1%) due to insufficient residual serum sample.

### Reverse Transcriptase Polymerase Chain Reaction

Reverse transcriptase polymerase chain reaction for SARS-CoV-2 determination in NP swab samples was performed on a LightCycler 480 Instrument II (Roche Diagnostics, Rotkreuz, Switzerland) using the LightMix Modular SARS-CoV E-gene set (for samples originating from Clinique Saint-Luc Bouge) and on the GeneXpert instrument (Cepheid, California, USA) using the Xpress SARS-CoV-2 assay targeting N2 and E genes (for samples originating from Clinique Saint-Luc Bouge and Clinique Saint-Pierre Ottignies). Cycle threshold (Ct) values obtained by RT-PCR were used as a proxy for the viral load.

### Statistical Analyses

Descriptive statistics were used to analyze the data. Smoothing splines with four knots were used to estimate the time kinetics curves using all longitudinal samples from the study population. Difference between disease severity per time intervals (i.e. < 3 days, 4 to 10 days, 11 to 20 days and > 20 days) and difference between time intervals per severity was assessed using an ordinary two-way ANOVA with Šidák’s multiple comparison tests with individual variances computed for each comparison. In the subpopulation A, antigen results have been used to define cut-offs for severity. Receiver operating characteristics (ROC) curve for antigen assays were performed and the corresponding area under the curves (AUC) was calculated. The Youden index was used to determine the optimal severity cut-off. Additionally, a positive likelihood ratio (sensitivity/1-specificity) was calculated based on the severity cut-off for prediction of severe forms of COVID-19.

In the subpopulation B, the comparison of RT-PCR and antigen results at the time of diagnosis (i.e. at the time of the RT-PCR), according to disease severity was tested using an ordinary one-way ANOVA. A Tukey multiple testing comparison was applied with multiplicity adjusted p value. Antigen and SARS-CoV-2 Spike IgG values were log-transformed. ROC curves were also performed and the Youden index was used to determine the optimal positivity cut-offs using subpopulation B. Additionally, clinical sensitivity was determined and represented the proportion of SARS-CoV-2 positive samples by antigen tests initially categorized as positive by RT-PCR. Clinical specificity was defined as the proportion of samples identified as negative by the antigen tests initially categorized as negative by RT-PCR. According to the Centers for Disease Control and Prevention (CDC), Ct values < 33 were considered as a surrogate of contagiousness and only these results were reported as true positive in this study.(15, 21)

Antigen level comparison between patients with or without SARS-CoV-2 Spike IgG response has been performed using a Welch’s t-test. Pearson’s correlation coefficients were used to investigate the correlation between RT-PCR, SARS-CoV-2 Spike IgG and N antigen results and between the two antigen assays.

Data analysis was performed using GraphPad Prism software (version 9.2.0, California, USA) and MedCalc software (version 14.8.1, Ostend, Belgium). P < 0.05 was used as a significance level. Our study fulfilled the Ethical principles of the Declaration of Helsinki.

## Results

### Kinetics of the N Antigen in Serum and Determination of Severity Cut-offs

The kinetics of the N antigen in the studied population is presented in **Figure 2** for both the Simoa and the iFlash assay. The peak antigen response was observed at day 7 in severe patients using both assays. Afterwards, a decline was observed up to day 20. In non-severe patients, the antigen response corresponded to a plateau phase that slowly decreased over time. The difference of kinetics between severe and non-severe patients was more distinguishable using the Simoa assay (**Figure 2**). Using ROC curves analyses on samples collected in patients from day 2 to day 14 (subpopulation B), the optimal cut-off to identify severe patients have been found to be 5,043 pg/mL for the Simoa (AUC = 0.927 [95%CI: 0.847-1.00], sensitivity = 84.6% [95%CI: 57.8-97.3], specificity = 97.2% [95%CI: 90.3-99.5, positive likelihood ratio = 30.0] and 313.8 COI for the iFlash (AUC = 0.895 [95%CI: 0.806-0.984], sensitivity = 76.9% [95%CI: 49.7-91.8], specificity = 93.0% [95%CI: 84.6-97.0], positive likelihood ratio = 10.9) (**Supplemental Figure 2**). The application of these severity cut-offs on kinetic models permitted to identify the best timing since symptom onset to identify severe patients (i.e. from 4 to 10 days) (**Figure 2 – red dotted lines**).

**Figure 2:**
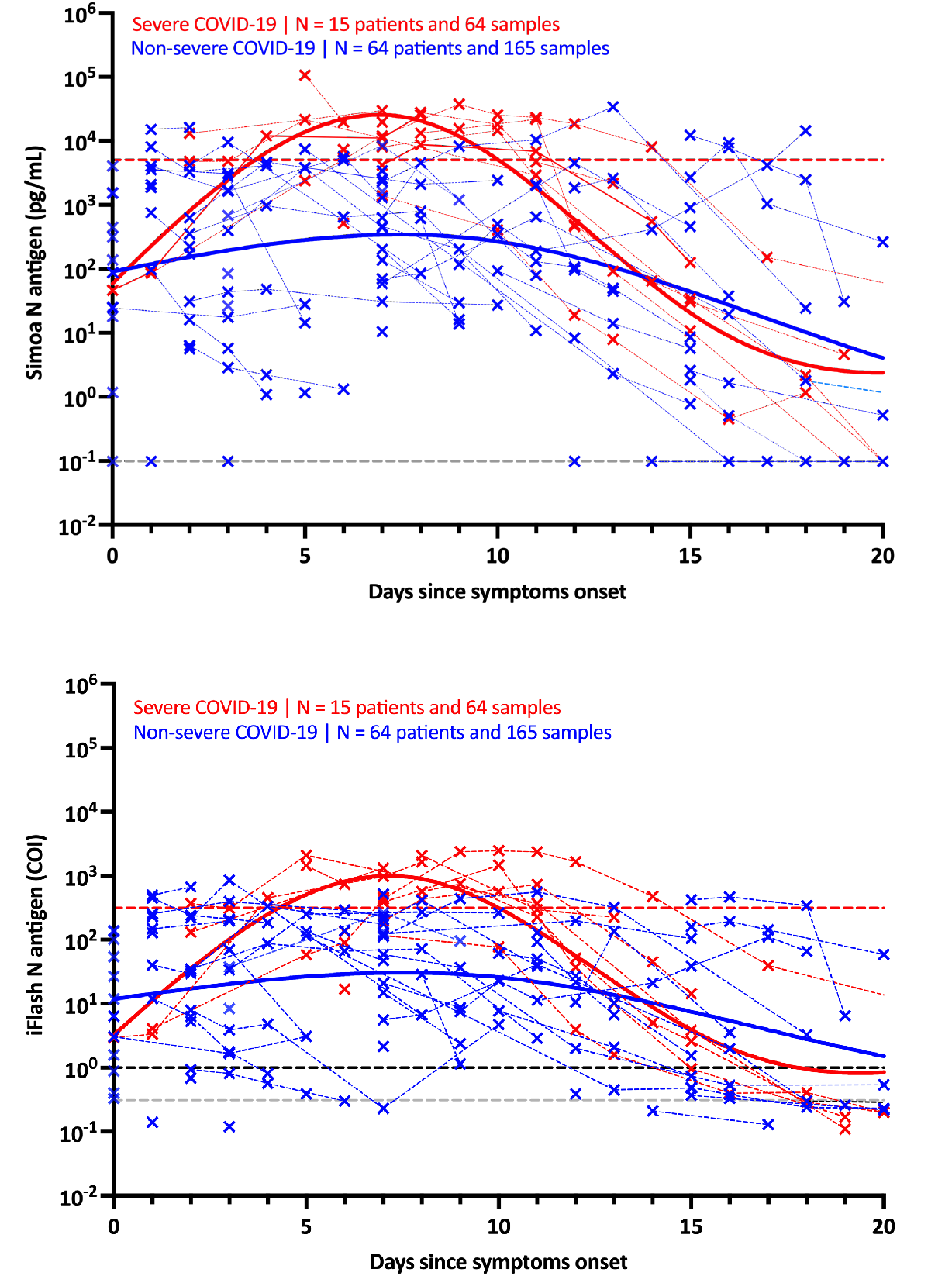
Kinetics of antigenemia since the onset of symptoms in non-severe and severe patients. The grey dotted lines correspond to the positivity cut-off of each antigen assay, as found by ROC curves analyses. The black dotted line corresponds to the positivity cut-off of the iFlash assay, as declared the manufacturer. The red dotted lines correspond to the severity cut-off of each antigen assay, as found by ROC curve analyses for the day 2 – day 14 window. Only patients with symptoms and negative for SARS-CoV-2 Spike IgG directed against the spike protein were included in this kinetics representation.

By splitting timing since symptom onset into categories (i.e. < 3 days, 4 to 10 days, 11 to 20 days and > 20 days), the antigen level in severe patients observed between days 4 and 10 was significantly higher compared to non-severe patients, whatever the time category. These observations were similar for both antigen assays (**Figure 3)**. Using the Simoa assay, the sensitivity of antigen in serum up to 10 days since symptom onset was 100% in severe patients and ranged from 91.3% (< 3 days) to 100% (from 4 to 10 days) in non-severe patients (global sensitivity = 96.9%). Between days 11 and 20, the sensitivities decreased to 88.5% and 86.5% and after 20 days waned at 75.0% and 35.3% in severe and non-severe patients (**Figure 3**). Using the iFlash assay with the optimized positivity cut-off of 0.31 COI (see section below), the sensitivity of antigen in serum up to 10 days since symptom onset was 100% in severe patients and ranged from 93.5% (< 3 days) to 96.0% (from 4 to 10 days) in non-severe patients (global sensitivity = 96.2%). Between days 11 and 20, the sensitivities decreased at 80.8% and 86.5% and after 20 days dampened at 75.0% and 23.5% in severe and non-severe patients.

**Figure 3:**
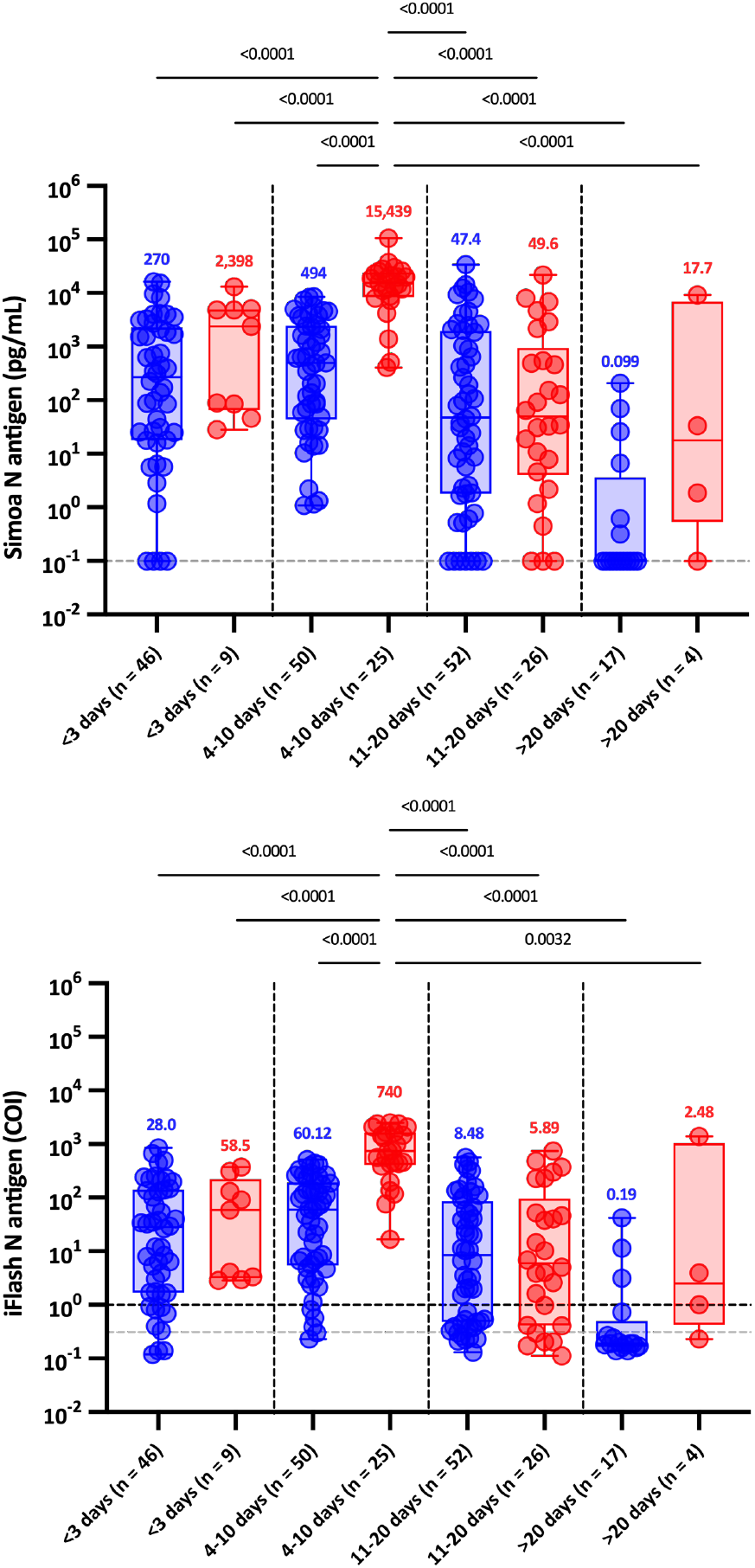
Levels of serum antigen according to the delay since the onset of symptoms (< 3, 4-10, 11-20, and > 20 days) and to severity. Blue dots correspond to non-severe patients and red dots to severe patients.The grey dotted lines correspond to the positivity cut-off of each antigen assay, as found by ROC curves analyses. The black dotted line corresponds to the positivity cut-off of the iFlash assay, as declared the manufacturer. Medians are represented on top of each whisker box.

These two antigen assays showed a highly significant correlation with a Pearson r of 0.96 (p < 0.0001) (**Supplemental Figure 3**). Among the 243 samples, only 12 (4.9%) were discordant. Four results were positive on the iFlash assay (range: 0.33-0.54 COI) but negative on the Simoa and 8 were negative on the iFlash assay but positive on the Simoa (range: 0.32-30.9 pg/mL).

### Clinical Performance of SARS-CoV-2 Antigen Assays in Serum Compared to RT-PCR from Nasopharyngeal Swabs

The mean Ct values obtained by RT-PCR were significantly lower in case of serum antigen positivity using the two antigen assays (p < 0.0001) (**Figure 4 – panel A**). The clinical sensitivity was 100% and the clinical specificity was 92.3% for both antigen assays (**Table 1**). Highly significant correlations were found between RT-PCR results and antigen levels (Pearson r of −0.54 [iFlash] to −0.61 [Simoa]; p < 0.0001) (**Figure 4 – panel B and C**).

**Figure 4:**
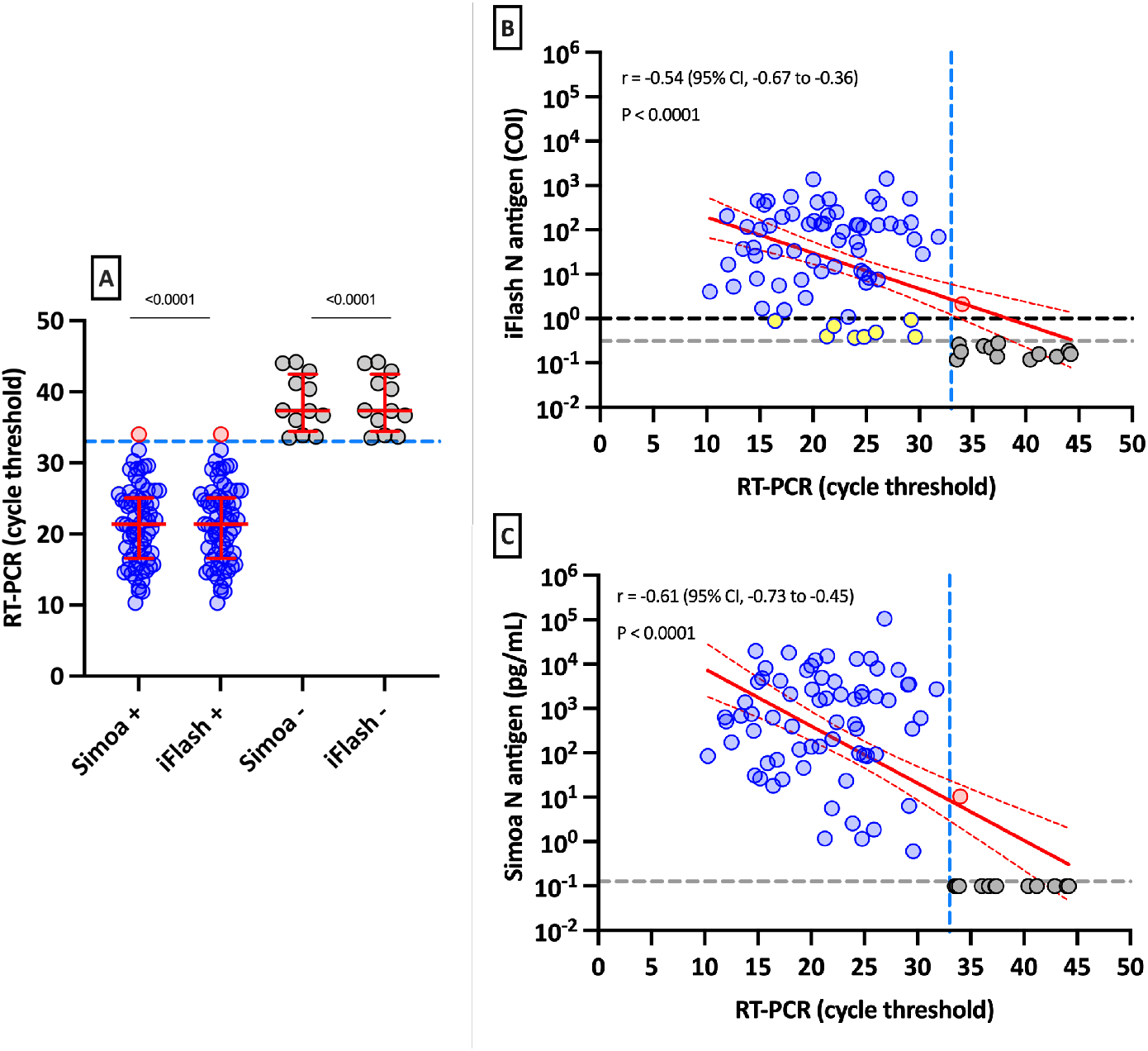
**[A] Positive and negative antigen results in serum according to RT-PCR cycle threshold values in nasopharyngeal samples.** The blue dotted line corresponds to a cycle threshold of 33. **[B-C] Linear regression of cycle threshold results obtained by RT-PCR versus antigenemia measured on the iFlash and the Simoa**. The grey dotted line on the Y-axis of the B and C panels represents positivity cut-off for antigen positivity according to the ROC curve. The black dotted line on the Y-axis of the B panel represents the cut-off for the iFlash N antigen as provided by the manufacturer. The blue dotted line on the X-axis corresponds to a cycle threshold of 33. Samples in yellow represent samples which become positive with the cut-off determined by the ROC curve as compared to the cut-off of the manufacturer. The sample is red represents a sample which is positive with both antigen assays and above a cycle threshold of 33 with the RT-PCR.

Based on the Youden index recommended values, the following positivity cut-offs were found for the iFlash and the Simoa assays: > 0.310 COI and > 0.099 pg/mL. The application of these cut-offs on the pre-pandemic cohort resulted in specificities of 95.8% (95%CI: 88.1-99.1; 3 false positive results) and 98.6% (95%CI: 92.4-99.9; 1 false positive result), respectively (**Table 1**). False positive pre-pandemic samples were different between the iFlash and the Simoa assays.

Antigen assays were also confronted to RT-PCR for determination of disease severity at the time of diagnosis (i.e. at the time of the RT-PCR). Considering NP RT-PCR, the mean Ct value of asymptomatic patients was significantly higher compared to severe patients (p = 0.0037). No significant difference was observed between mild, moderate, and severe patients based on NP RT-PCR (**Figure 5**). Serum antigen assays showed higher antigen levels in severe patients (**Figure 5**). The difference was statistically significant between severe patients and asymptomatic or moderate patients. Moderate patients also had significantly higher antigen levels compared to asymptomatic subjects.

**Figure 5:**
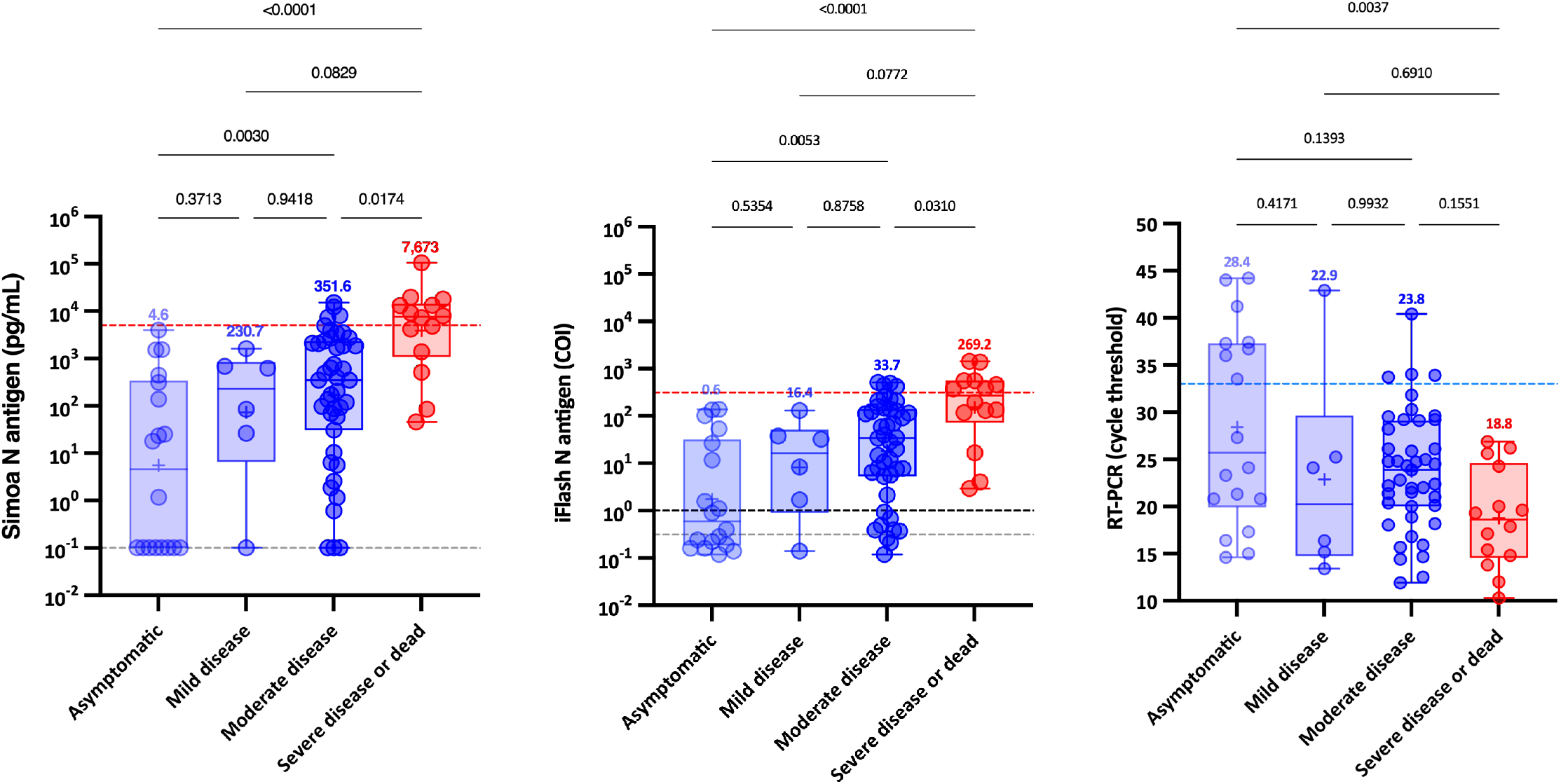
Antigenemia and RT-PCR results according to the WHO clinical progression scale on samples obtained on the day of diagnosis, i.e. within 12 hours since the RT-PCR. The blue dotted lines correspond to a cycle threshold of 33. The red dotted line corresponds to the severity cut-off, as determined by the ROC curve analyze. The grey dotted lines correspond to the positivity cut-off of each antigen assay, as found by ROC curves analyses. The black dotted line corresponds to the positivity cut-off of the iFlash assay, as declared the manufacturer. Medians are represented on top of each whisker box.

### Correlation of the Serological Status with SARS-CoV-2 Antigen Assays in Serum and RT-PCR from Nasopharyngeal Swabs

For the 77 samples for which the serological status has been obtained at the time of diagnosis (i.e. at the time of the RT-PCR), 18 had positive SARS-CoV-2 Spike IgG (23.4%) and 59 were considered negative (76.6%). The mean number of Ct obtained by RT-PCR was significantly lower in patients negative for SARS-CoV-2 Spike IgG (p < 0.0001) and most negative IgG patients (94.9%) presented a Ct value < 33 (**Supplemental Figure 4 – panel A**). A positive correlation between SARS-CoV-2 Spike IgG and RT-PCR results was found (r = 0.67; p < 0.0001) (**Supplemental Figure 4 – panel B**).

The mean concentration of serum antigens was significantly lower in SARS-CoV-2 Spike IgG positive samples (P value < 0.0001) (**Supplemental Figure 5 – panel A**) and negative correlations between SARS-CoV-2 Spike IgG and antigen results were also identified (Pearson r of −0.60 [iFlash] to −0.66 [Simoa]; p < 0.0001) (**Supplemental Figure 5 – panel B**). Interestingly, patients with a Ct value > 33 along with negative antigen in serum all had positive SARS-CoV-2 Spike IgG (n = 12) (**Figure 4**). Contrariwise, most patients (96.6%) with negative SARS-CoV-2 Spike IgG were positive for antigen in serum using both assays (**Supplemental Figure 5 – panel A**).

## Discussion

To date, few studies have investigated the interest of detecting blood antigen of patients with SARS-CoV-2 infection.(4-6, 15, 22, 23) In this study which included 90 patients for whom 243 blood samples were obtained at different times since symptom onset, we evaluated the kinetics of the SARS-CoV-2 N antigen in serum from infected patients using two antigen assays, i.e. the Simoa and the iFlash assays. The disease severity was evaluated using to the WHO clinical progression scale(18).

Like previous studies investigating the possible prognostic value of antigen testing in serum samples,(5, 6, 24-26) we evaluate whether these antigen assays could help in identifying patients more at risk of developing a severe form of COVID-19. These studies revealed that a N antigen determination at the time of diagnosis, especially within the first week of symptom onset, may have potential value in triaging patients for higher-level care.(23, 26) Higher concentrations of N antigen were observed in more severe patients (5, 6, 23-26) as well as positive correlations with inflammatory biomarker levels (i.e. CRP or IL-6).(24, 25, 27) Our study confirms that severe patients exhibit higher N antigen levels compared to non-severe patients both at the time of diagnosis (i.e. at the time the NP RT-PCR was performed – **Figure 5**) and during the first days since the onset of symptoms (**Figure 2 and 3**). Difference between severe and non-severe patients was especially noticeable between days 4 and 10. We also estimated that cut-offs for identifying patients more at risk to be severe were 5,043 pg/mL and 313.8 COI on the Simoa and the iFlash assays. The likelihood ratio for these cut-offs were 30.0 and 10.9 which reflects their good capacity to distinguish severe from non-severe patients during the day 2 to day 14 timeframe. The likelihood ratio was lower using the iFlash assay and the larger range of antigen concentration of the Simoa technology (from 0.099 to 100,000 pg/mL) might explain why the Simoa may be a better predictor of severe outcomes compared to the iFlash, even if a high correlation was found between the two techniques (**Supplemental Figure 2**). Compared to N antigen in serum, Ct values of RT-PCR failed to predict COVID-19 severity **(Figure 5**), an observation which is similar to the one of Wang et al.(23)

Various assays (Simoa, Lumipulse G, Quantigene, and Biohit healthcare assays) revealed a specificity ranging from 98.4% to 100% on pre-pandemic samples.(5, 15, 25, 26) Ogata *et al*. were the first to report on the Simoa technology and found a specificity of 82.4%.(6) The method they used was, however, home-made and do not represent the current performance of the Simoa assay. As a matter of fact, Shan *et al*. found a specificity of 100% using the commercial kit from Quanterix on the Simoa.(5) In our evaluation, specificities of the iFlash and Simoa were 95.8% and 98.6% and were therefore consistent with the data from the literature.

The clinical performance of these antigen assays was also directly compared to RT-PCR performed on NP swabs.(5, 15, 25, 26) The clinical performance is directly related to the design of published studies. If considering studies that included patients that developed symptoms up to maximum 2 weeks, the clinical sensitivity ranged from 85.2% to 93.0% (15, 23, 26) and increased to 94.2% to 100% with samples collected within the first days since symptoms.(15, 24) However, the clinical sensitivity significantly decreased after 2 weeks (43.2% to 74.5%) (5, 15, 23, 26) and > 4 weeks since symptoms (from 0% to 34.2%.(23, 28) Longitudinal monitoring of antigen concentrations more than 2 weeks after the onset of symptoms is therefore not likely to be helpful in predicting the outcome or response to therapy.(23) Six studies found moderate clinical sensitivities ranging from 41.0% to 74.0%.(6, 22, 24, 25, 27, 29) This lower performance is explained by the design of these studies. Indeed, the timing since symptoms was either not disclosed (22, 24, 25, 29) or was long (i.e. up to 30 days since symptoms), decreasing the probability of having positive samples knowing the kinetic of the SARS-CoV-2 N antigen in blood.(6, 27) Given that the peak of N antigen is reached after 7 days, as for the viral load in NP samples,(30) and that a continuous decline is observed afterwards, the timing since symptoms is a paramount information for the evaluation of antigenemia (**Figure 2**). In our evaluation, we confirmed that the clinical sensitivity of N antigen as compared to RT-PCR was excellent (96.2% for iFlash and 96.9% for Simoa) in samples collected earlier since symptom onset (i.e. < 10 days), especially in more severe patients (i.e. 100% for both assays) (**Figure 3**), as previously reported.(23, 26)

When considering serum samples collected < 12 hours from NP RT-PCR (mean delta time = 23 minutes, **Supplemental Figure 3**), the clinical sensitivity for Ct < 33 was 100% for both tests using the optimized cut-offs. Indeed, patients with low viral load (i.e. Ct > 33) have a very low risk of being infective and capable of transmitting the virus. Therefore, the NP RT-PCR positivity in these subjects may be attributable to residual low viral load.(31, 32) If considering each Ct value as true positive result, the clinical sensitivity decreased to 85.2% (69/81). The adjunction of a SARS-CoV-2 Spike IgG test identified that these 12 samples (all in the non-severe patients’ group) with a high Ct value (i.e. Ct > 33) but negative N antigen, all had positive antibodies (i.e. levels of IgG antibodies > 924 ng/mL). Therefore, this explains why the serum antigen results were negative, as these patients presented after seroconversion. Wang *et al*. also found that negative N antigen in COVID-19 patients corresponded to higher Ct values (38.2 = median; IQR = 37.4-39.2).(23)

Interestingly, patients who exhibited a level of SARS-CoV-2 Spike IgG above the positivity cut-off showed statistically significant higher Ct values and lower serum antigen levels (**Supplemental Figures 4 and 5**). As expected from previous studies in COVID-19 subjects, the level of antibodies started to increase approximately 10 days after the onset of symptoms. It also correlates with the waning of N antigen levels as observed in **Supplemental Figure 6** and in another study.(23) These data support the concept that antigenemia may be a prognostic marker of severity to identify patients more likely to require intensive care in the first few days after the onset of symptoms.

The clinical specificity of N antigen assays ranged from 68.0% to 99.8% (mean clinical specificity = 90.9%),(6, 22, 23, 26, 28, 29) and were in line with the results we reported in this study (92.3%) (**Table 1**). This implies that some patients have positive N antigen in blood but negative NP RT-PCR. It should be noted that this could be a false negative NP RT-PCR from a true infected patient. In the study of Le Hingrat *et al*., 8 out of 12 patients with a negative RT-PCR result had detectable N antigen in blood. In 6 of these 8 patients (i.e. 75%), RT-PCR performed on lower respiratory tract samples turned out positive showing that the virus may have migrated from the nasopharyngeal sphere to the lower respiratory tract.(15) Additionally, Su *et al*. found that 16 (32.2%) samples from 50 COVID-19 negative patients by pharyngeal RT-PCR were positives for the presence of N antigen, including two severe COVID-19 patients.(22) The detection of blood antigen can therefore be a valuable alternative to RT-PCR, especially in case of negative results. In our study, we identified a patient with positive N antigen in serum (both on iFlash and Simoa, low positive sample: 2.14 COI and 10.5 pg/mL) but with a Ct value > 33 (i.e. Ct of 34) and positive for SARS-CoV-2 Spike IgG antibodies (**Figure 4**).

In a previous study, the application of ROC curve adapted cut-offs (1.85 to 10 pg/mL) allowed to significantly increase the clinical sensitivity (from 76 to 92%) with a limited impact on the specificity (from 100% to 96.84%).(29) In our study, we increased the clinical performance of the iFlash (1.0 to 0.31 COI). This approach has also been successfully used for SARS-CoV-2 antibody assays.(33)

## Conclusion

Sensitive N antigen detection in serum provides a valuable new marker for COVID-19 diagnosis, only requiring a blood draw, that is scalable in all clinical laboratories. It allows potential new developments to design rapid antigen blood tests or combined ELISA assays, detecting both antigens and antibodies. Measuring antigen in blood present several advantages including a more standardized process of obtaining blood samples compared to RT-PCR with possible development to dry blood spots sampling strategies. Importantly, antigenemia, when assessed during the first 2 weeks since the onset of symptoms, may help in identifying patients at risk of developing severe COVID-19. The severity cut-offs proposed in our investigation need to be confirmed at subsequent studies, but it already supports the rational that compared to the NP RT-PCR, the detection of N antigen in blood allows a better discrimination of severe cases from non-severe ones. In addition, these techniques are at least as accurate than NP RT-PCR for diagnosing SARS-CoV-2 infection, giving us the possibility to develop more convenient testing strategies for patients. It may finally facilitate patient triage to optimize better intensive care utilization.

## Data Availability

All data produced in the present study are available upon reasonable request to the corresponding author

## Acknowledgment

The authors would like to thank the technical teams from the laboratories of QUALIblood, Clinique Saint-Luc Bouge and Clinique Saint-Pierre Ottignies for collecting the samples and performing the analyses.

## Author contributions

Conceptualization: JFA – JBA – JDO; Data curation: JFA – JBA – CDA – CGI – CEU – JDO; Formal analysis: JFA – JBA – CDA – GWI – GRO – GSO – JDO; Funding acquisition: JDO; Investigation: JFA – CEU – JDO; Methodology: JFA-JDO; Project administration: MEL – JMD – JDO; Resources: MEL – CEU – JDM – JDO; Supervision: JDO; Validation: JFA – JDO; Visualization: JFA – GWI – GRO – JDO; Writing – original draft: JFA – CDA – JDO; Writing – review & editing: JFA – JBA – CDA – CGI – GWI – GRO – GSO – MEL – CEU – JMD – JDO

## Data availability statement

The data that support the findings of this study are available from the corresponding author JDO, upon reasonable request.

---

**supplementary Table 1:** Demographic data of the included population.

| **Demography** |  |
| --- | --- |
| **Participants (n)** | **90** |
| **Age (median [min-max])** | **77 [20-97]** |
| **Females (n [%])** | **44 [48.9%]** |
| *Age (median [min-max])* | *78 [20-97]* |
| **Males (n [%])** | **46 [51.1%]** |
| *Age (median [min-max])* | *77 [33-94]* |
| **Clinical status** |  |
| **Asymptomatic (n [%])<sup>1</sup>** | **11 [12.2%]** |
| **Symptomatic (n [%])** | **79 [83.3%]** |
| *Mild disease (n [%])<sup>2</sup>* | *17 [18.9%]* |
| *Moderate disease (n [%])<sup>3</sup>* | *47 [52.2%]* |
| *Severe disease (n [%])<sup>4</sup>* | *15 [16.7%]* |

<sup>1</sup> WHO clinical progression scale of 1; <sup>2</sup> WHO clinical progression scale of 2-3; <sup>3</sup> WHO clinical progression scale of 4-5; <sup>4</sup> WHO clinical progression scale of 6-10

**supplemental Figure 1:**
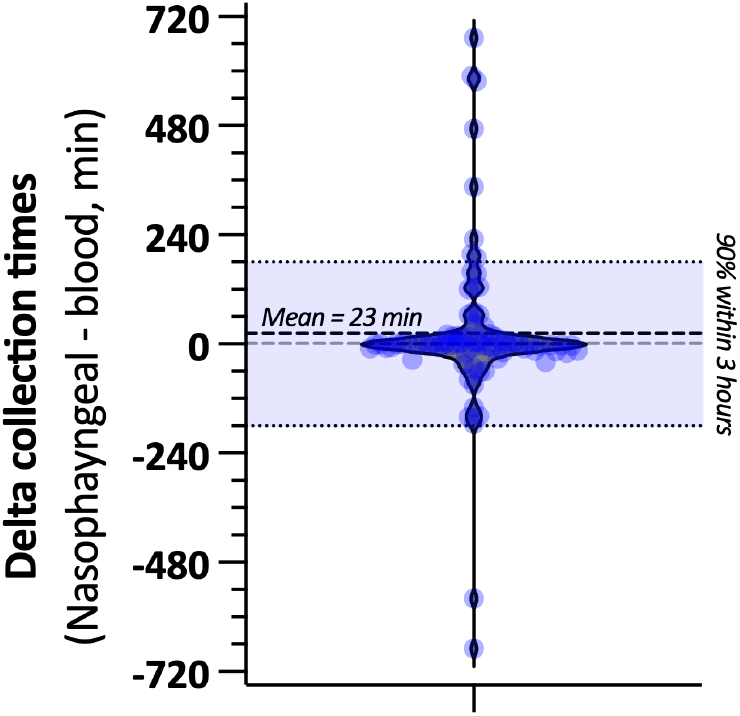
Timing between the collection of NP and blood samplings, in minutes. For 41 samples, the NP was collected before the blood sample. For the other 40 samples, the blood sample was collected before the NP swab. The median delta collection time was 1 minute (10^th^ − 90^th^ percentile: −74 to 183 minutes), and the mean delta time was 23 minutes (standard deviation: 179 minutes)

**supplemental Figure 2:**
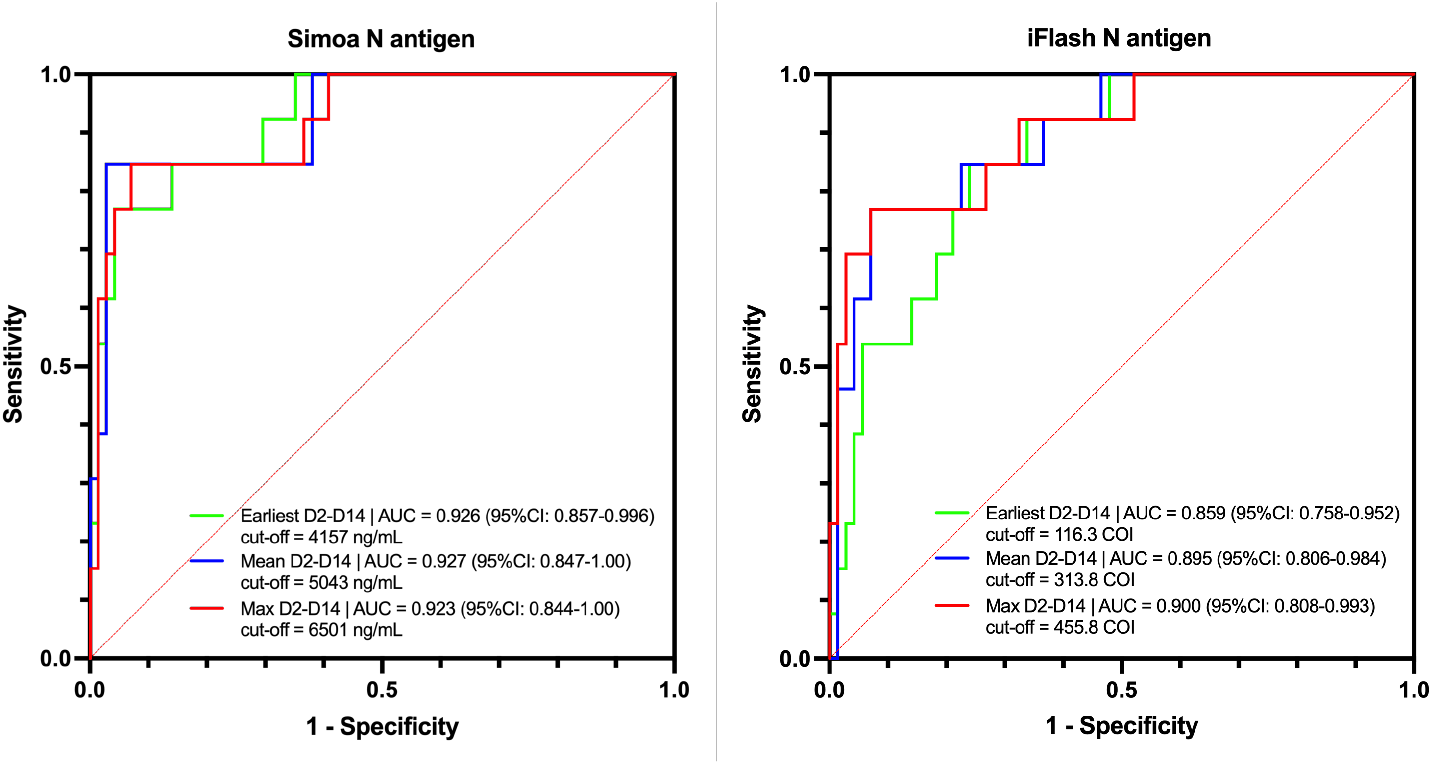
ROC curves for cut-off determination according to different samples inclusion settings. Three different inclusion settings were used: a) earliest antigenemia value from symptom onset within the day 2 to day 14 window for a particular patient, b) mean of all antigenemia values of samples collected within the day 2 to day 14 window for a particular patient and c) the maximal antigenemia value obtained within the day 2 to day 14 window. For all these analyses, 84 samples were included (n = 71 for non-severe patients and n = 13 for severe patients). These analyses were performed for the Simoa and the iFlash assays.

**supplemental Figure 3:**
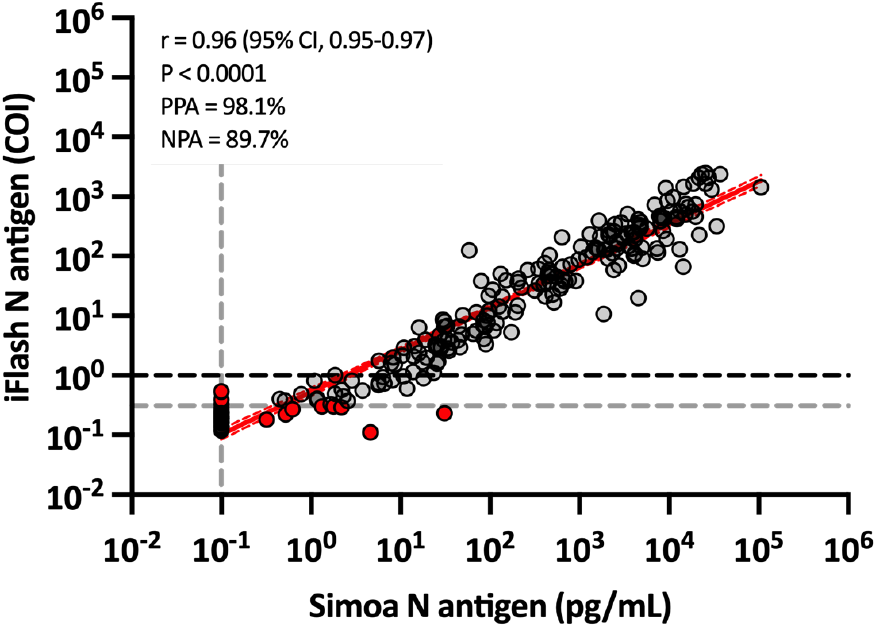
Correlation between the Simoa and the iFlash N antigen assays. The grey dotted lines represent the cut-off determined by the ROC curves and the black dotted line on the iFlash axis represent the cut-off of the manufacturer. *Abbreviations: NPA, negative percentage agreement; PPA, positive percentage agreement*.

**Supplemental Figure 4:**
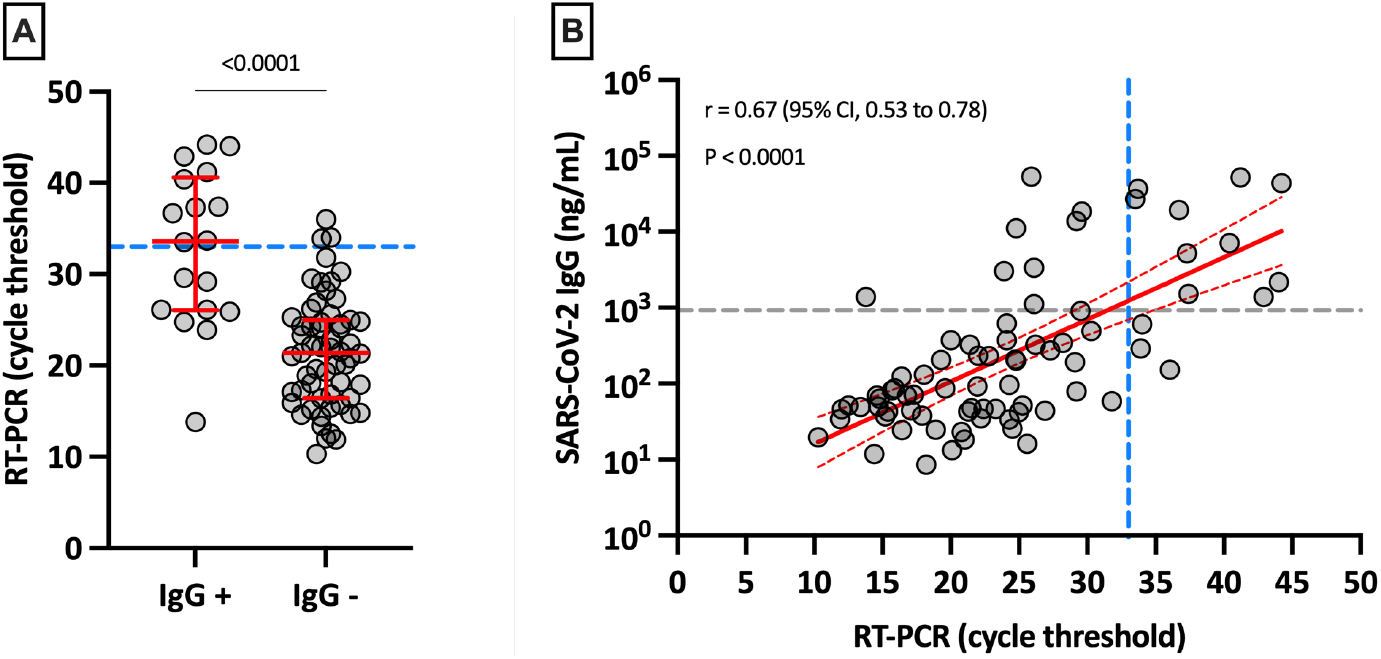
[A] Positive and negative SARS-CoV-2 Spike IgG results in serum according to RT-PCR cycle threshold results. The blue dotted line corresponds to a cycle threshold of 33. **[B] Linear regression of Ct results obtained by RT-PCR versus the amount of SARS-CoV- 2 Spike IgG obtained on the Simoa.** The grey dotted line on the Y-axis represents positivity cut-off for IgG directed against the spike protein. The blue dotted line on the X-axis corresponds to a cycle threshold of 33.

**supplemental Figure 5:**
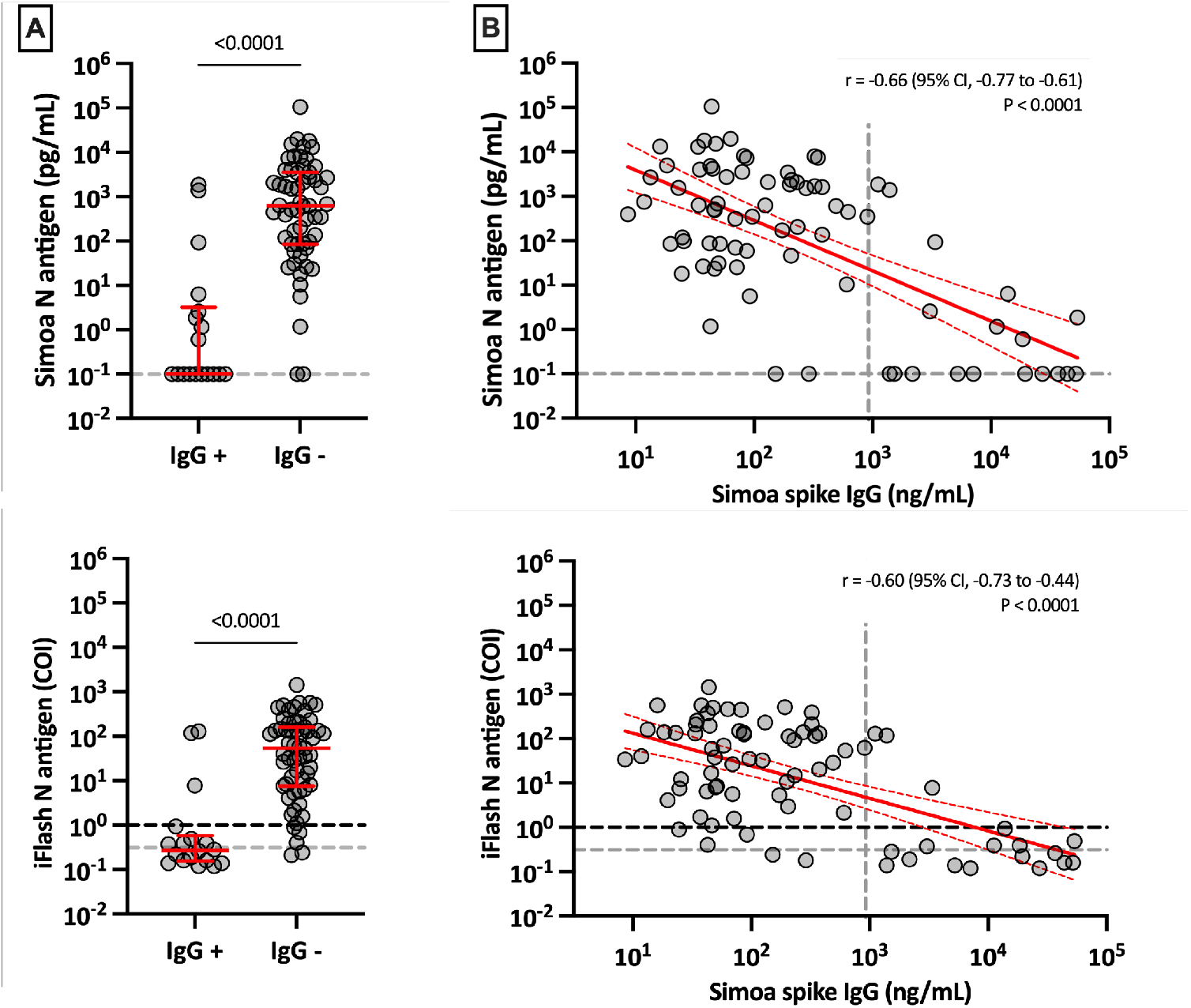
[A] Positive and negative SARS-CoV-2 Spike IgG results in serum according to antigenemia. The grey dotted lines on the Y-axis correspond to the positivity cut-off of each antigen assay, as found by ROC curves analyses. The black dotted line on the Y-axis of the iFlash panel represents the cut-off of the manufacturer. **[B] Linear regression of antigenemia obtained with the two antigen assays versus the amount of SARS-CoV-2 Spike IgG**. The grey dotted line on the X-axis represents positivity cut-off for IgG directed against the spike protein.

**supplemental Figure 6:**
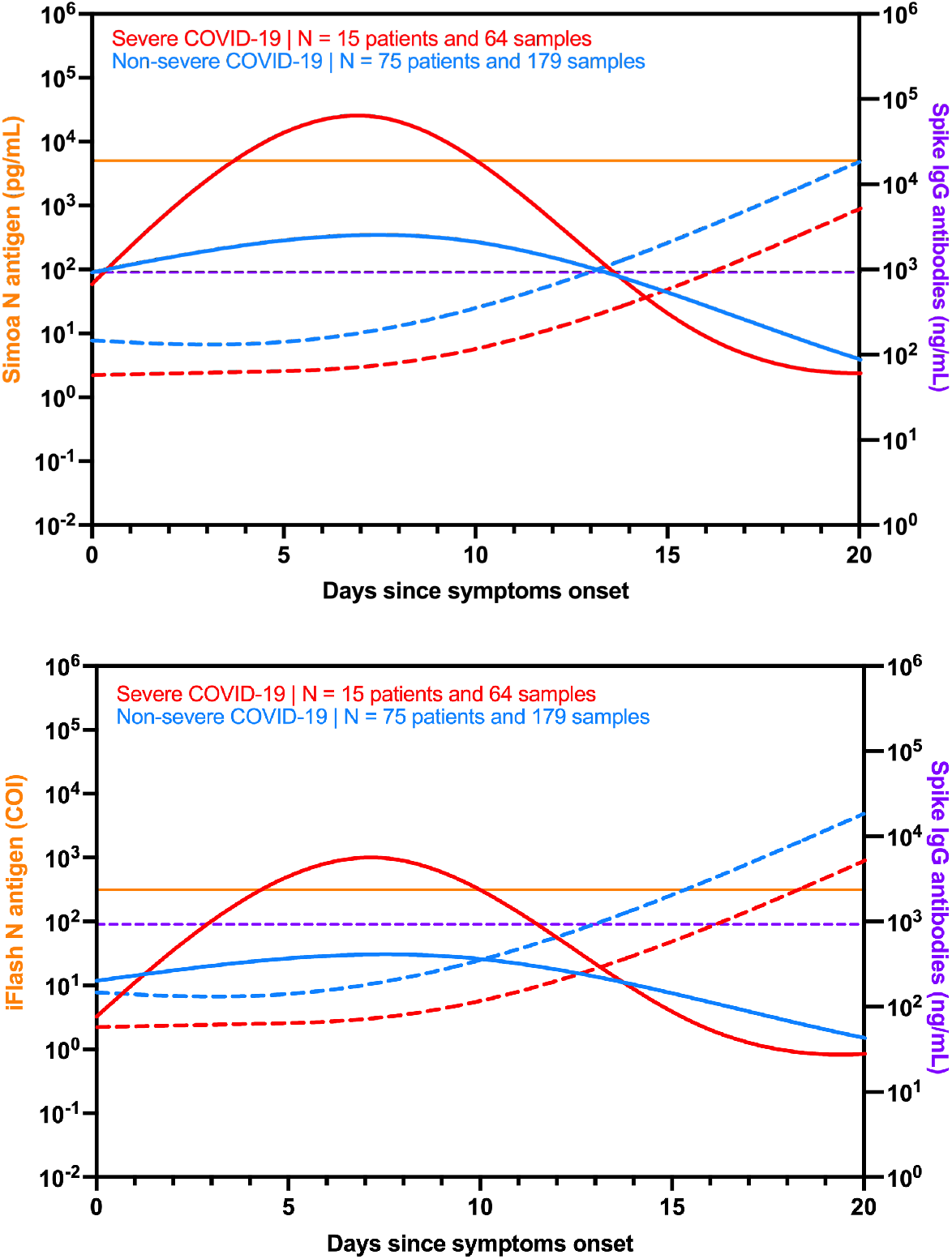
Kinetics of antigenemia and SARS-CoV-2 Spike IgG antibodies since the onset of symptoms in non-severe and severe patients determined according to the WHO clinical progression scale.(18) The continuous orange lines correspond to the severity cut-off of each antigen assay, as found by ROC curves analyses. The purple dotted lines correspond to the positivity cut-off of the SARS-CoV-2 Spike IgG assay. The continuous red and blue lines correspond to antigen kinetics in severe and non-severe patients. The dotted red and blue lines correspond to the kinetics of SARS-CoV-2 Spike IgG in severe and non-severe patients. Only patients with symptoms and negative for IgG directed against the Spike protein were included in this representation.

## STARD checklist

STARD checklist:
| Section & Topic | No | Item | Reported on page # |
| --- | --- | --- | --- |
| **TITLE OR ABSTRACT** |  |  |  |
|  | 1 | Identification as a study of diagnostic accuracy using at least one measure of accuracy (such as sensitivity, specificity, predictive values, or AUC) | 3 |
| **ABSTRACT** |  |  |  |
|  | 2 | Structured summary of study design, methods, results, and conclusions (for specific guidance, see STARD for Abstracts) | 3 |
| **INTRODUCTION** |  |  |  |
|  | 3 | Scientific and clinical background, including the intended use and clinical role of the index test | 4-5 |
|  | 4 | Study objectives and hypotheses | 5 |
| **METHODS** |  |  |  |
| *Study design* | 5 | Whether data collection was planned before the index test and reference standard were performed (prospective study) or after (retrospective study) | 5 |
| *Participants* | 6 | Eligibility criteria | 5 |
|  | 7 | On what basis potentially eligible participants were identified (such as symptoms, results from previous tests, inclusion in registry) | 5 |
|  | 8 | Where and when potentially eligible participants were identified (setting, location and dates) | 5 |
|  | 9 | Whether participants formed a consecutive, random or convenience series | 5 |
| *Test methods* | 10a | Index test, in sufficient detail to allow replication | 6-8 |
|  | 10b | Reference standard, in sufficient detail to allow replication | 6-8 |
|  | 11 | Rationale for choosing the reference standard (if alternatives exist) | 3 |
|  | 12a | Definition of and rationale for test positivity cut-offs or result categories of the index test, distinguishing pre-specified from exploratory | 8-9 |
|  | 12b | Definition of and rationale for test positivity cut-offs or result categories of the reference standard, distinguishing pre-specified from exploratory | 8-9 |
|  | 13a | Whether clinical information and reference standard results were available to the performers/readers of the index test | 5 |
|  | 13b | Whether clinical information and index test results were available to the assessors of the reference standard | 5 |
| *Analysis* | 14 | Methods for estimating or comparing measures of diagnostic accuracy | 8-9 |
|  | 15 | How indeterminate index test or reference standard results were handled | 8-9 |
|  | 16 | How missing data on the index test and reference standard were handled | 8-9 |
|  | 17 | Any analyses of variability in diagnostic accuracy, distinguishing pre-specified from exploratory | 8-9 |
|  | 18 | Intended sample size and how it was determined | / |
| **RESULTS** |  |  |  |
| *Participants* | 19 | Flow of participants, using a diagram | 5 and Figure 1 |
|  | 20 | Baseline demographic and clinical characteristics of participants | 6 and Supplemental Table 1 |
|  | 21a | Distribution of severity of disease in those with the target condition | 6 |
|  | 21b | Distribution of alternative diagnoses in those without the target condition | 9-11 |
|  | 22 | Time interval and any clinical interventions between index test and reference standard | 5 and Supplemental Figure 1 |
| *Test results* | **23** | Cross tabulation of the index test results (or their distribution) by the results of the reference standard | 9-12 |
|  | **24** | Estimates of diagnostic accuracy and their precision (such as 95% confidence intervals) | 9-12 |
|  | **25** | Any adverse events from performing the index test or the reference standard | / |
| **DISCUSSION** |  |  |  |
|  | **26** | Study limitations, including sources of potential bias, statistical uncertainty, and generalisability | 12-15 |
|  | **27** | Implications for practice, including the intended use and clinical role of the index test | 15 |
| **OTHER INFORMATION** |  |  |  |
|  | **28** | Registration number and name of registry | / |
|  | **29** | Where the full study protocol can be accessed | 6 |
|  | **30** | Sources of funding and other support; role of funders | 1 |

## References

1. Bohn MK, Lippi G, Horvath A, Sethi S, Koch D, Ferrari M, Wang CB, Mancini N, Steele S, Adeli K. 2020. Molecular, serological, and biochemical diagnosis and monitoring of COVID-19: IFCC taskforce evaluation of the latest evidence. Clin Chem Lab Med 58:1037–1052.

2. Quanterix. 2021. Simoa® SARS-CoV-2 N Protein Antigen Test — Instruction for use (IFU 0002). https://www.fda.gov/media/144929/download. Accessed

3. Favresse J, Gillot C, Oliveira M, Cadrobbi J, Elsen M, Eucher C, Laffineur K, Rosseels C, Van Eeckhoudt S, Nicolas JB, Morimont L, Dogne JM, Douxfils J. 2021. Head-to-Head Comparison of Rapid and Automated Antigen Detection Tests for the Diagnosis of SARS-CoV-2 Infection. J Clin Med 10.

4. Chen H, Li Z, Feng S, Richard-Greenblatt M, Hutson E, Andrianus S, Glaser LJ, Rodino KG, Qian J, Jayaraman D, Collman RG, Glascock A, Bushman FD, Lee JS, Cherry S, Fausto A, Weiss SR, Koo H, Corby PM, Oceguera A, O’Doherty U, Garfall AL, Vogl DT, Stadtmauer EA, Wang P. 2021. Femtomolar SARS-CoV-2 Antigen Detection Using the Microbubbling Digital Assay with Smartphone Readout Enables Antigen Burden Quantitation and Tracking. Clin Chem doi:10.1093/clinchem/hvab158.

5. Shan D, Johnson JM, Fernandes SC, Suib H, Hwang S, Wuelfing D, Mendes M, Holdridge M, Burke EM, Beauregard K, Zhang Y, Cleary M, Xu S, Yao X, Patel PP, Plavina T, Wilson DH, Chang L, Kaiser KM, Nattermann J, Schmidt SV, Latz E, Hrusovsky K, Mattoon D, Ball AJ. 2021. N-protein presents early in blood, dried blood and saliva during asymptomatic and symptomatic SARS-CoV-2 infection. Nat Commun 12:1931.

6. Ogata AF, Maley AM, Wu C, Gilboa T, Norman M, Lazarovits R, Mao CP, Newton G, Chang M, Nguyen K, Kamkaew M, Zhu Q, Gibson TE, Ryan ET, Charles RC, Marasco WA, Walt DR. 2020. Ultra-Sensitive Serial Profiling of SARS-CoV-2 Antigens and Antibodies in Plasma to Understand Disease Progression in COVID-19 Patients with Severe Disease. Clin Chem 66:1562–1572.

7. Mak GC, Lau SS, Wong KK, Chow NL, Lau CS, Lam ET, Chan RC, Tsang DN. 2020. Analytical sensitivity and clinical sensitivity of the three rapid antigen detection kits for detection of SARS-CoV-2 virus. J Clin Virol 133:104684.

8. Wang X, Yao H, Xu X, Zhang P, Zhang M, Shao J, Xiao Y, Wang H. 2020. Limits of Detection of 6 Approved RT-PCR Kits for the Novel SARS-Coronavirus-2 (SARS-CoV-2). Clin Chem 66:977–979.

9. Zou L, Ruan F, Huang M, Liang L, Huang H, Hong Z, Yu J, Kang M, Song Y, Xia J, Guo Q, Song T, He J, Yen HL, Peiris M, Wu J. 2020. SARS-CoV-2 Viral Load in Upper Respiratory Specimens of Infected Patients. N Engl J Med 382:1177–1179.

10. Han MS, Byun JH, Cho Y, Rim JH. 2021. RT-PCR for SARS-CoV-2: quantitative versus qualitative. Lancet Infect Dis 21:165.

11. Wolfel R, Corman VM, Guggemos W, Seilmaier M, Zange S, Muller MA, Niemeyer D, Jones TC, Vollmar P, Rothe C, Hoelscher M, Bleicker T, Brunink S, Schneider J, Ehmann R, Zwirglmaier K, Drosten C, Wendtner C. 2020. Virological assessment of hospitalized patients with COVID-2019. Nature 581:465–469.

12. Woloshin S, Patel N, Kesselheim AS. 2020. False Negative Tests for SARS-CoV-2 Infection - Challenges and Implications. N Engl J Med 383:e38.

13. Atyeo C, Fischinger S, Zohar T, Slein MD, Burke J, Loos C, McCulloch DJ, Newman KL, Wolf C, Yu J, Shuey K, Feldman J, Hauser BM, Caradonna T, Schmidt AG, Suscovich TJ, Linde C, Cai Y, Barouch D, Ryan ET, Charles RC, Lauffenburger D, Chu H, Alter G. 2020. Distinct Early Serological Signatures Track with SARS-CoV-2 Survival. Immunity 53:524–532 e4.

14. Sharma J, Rajput R, Bhatia M, Arora P, Sood V. 2021. Clinical Predictors of COVID-19 Severity and Mortality: A Perspective. Frontiers in Cellular and Infection Microbiology 11.

15. Hingrat QL, Visseaux B, Laouenan C, Tubiana S, Bouadma L, Yazdanpanah Y, Duval X, Burdet C, Ichou H, Damond F, Bertine M, Benmalek N, Choquet C, Timsit JF, Ghosn J, Charpentier C, Descamps D, Houhou-Fidouh N, French Covid cohort management committee C-Csg, members of the French Ccsg, member of the Co VCsgPi, Steering C, Co VCCC, Coordination, statistical a, Virological L, Biological C, Partners, Sponsor, Genetic. 2020. Detection of SARS-CoV-2 N-antigen in blood during acute COVID-19 provides a sensitive new marker and new testing alternatives. Clin Microbiol Infect doi:10.1016/j.cmi.2020.11.025.

16. Che XY, D. B, Zhao GP, Wang YD, Qiu LW, Hao W, Wang M, Qin PZ, Liu YF, Chan KH, Cheng VC, Yuen KY. 2006. A patient with asymptomatic severe acute respiratory syndrome (SARS) and antigenemia from the 2003-2004 community outbreak of SARS in Guangzhou, China. Clin Infect Dis 43:e1–5.

17. Huang C, Wang Y, Li X, Ren L, Zhao J, Hu Y, Zhang L, Fan G, Xu J, Gu X, Cheng Z, Yu T, Xia J, Wei Y, Wu W, Xie X, Yin W, Li H, Liu M, Xiao Y, Gao H, Guo L, Xie J, Wang G, Jiang R, Gao Z, Jin Q, Wang J, Cao B. 2020. Clinical features of patients infected with 2019 novel coronavirus in Wuhan, China. Lancet 395:497–506.

18. Marshall JC, Murthy S, Diaz J, Adhikari NK, Angus DC, Arabi YM, Baillie K, Bauer M, Berry S, Blackwood B, Bonten M, Bozza F, Brunkhorst F, Cheng A, Clarke M, Dat VQ, de Jong M, Denholm J, Derde L, Dunning J, Feng X, Fletcher T, Foster N, Fowler R, Gobat N, Gomersall C, Gordon A, Glueck T, Harhay M, Hodgson C, Horby P, Kim Y, Kojan R, Kumar B, Laffey J, Malvey D, Martin-Loeches I, McArthur C, McAuley D, McBride S, McGuinness S, Merson L, Morpeth S, Needham D, Netea M, Oh M-D, Phyu S, Piva S, Qiu R, Salisu-Kabara H, et al. 2020. A minimal common outcome measure set for COVID-19 clinical research. The Lancet Infectious Diseases 20:e192–e197.

19. YHLO. 2020. Instruction for Use — iFlash-2019-nCOV Antigen assay kit.

20. Quanterix. 2020. Instruction for Use — Simoa® SARS-CoV-2 Spike IgG Advantage Kit. DS-0521 01DS-0521.

21. Centers for Disease Control and Prevention. 2020. Common Investigation Protocol for Investigating Suspected SARS-CoV-2 Reinfection. https://www.cdc.gov/coronavirus/2019-ncov/php/reinfection.html. Accessed 29 Nov.

22. Su B, Yin J, Lin X, Zhang T, Yao X, Xu Y, Lu Y, Wang W, Liu K, Zhang J, Xie L, Jin R, Feng Y. 2021. Quantification of SARS-CoV-2 antigen levels in the blood of patients with COVID-19. Sci China Life Sci 64:1193–1196.

23. Wang H, Hogan CA, Verghese M, Solis D, Sibai M, Huang C, Roltgen K, Stevens BA, Yamamoto F, Sahoo MK, Zehnder J, Boyd SD, Pinsky BA. 2021. SARS-CoV-2 Nucleocapsid Plasma Antigen for Diagnosis and Monitoring of COVID-19. Clin Chem doi:10.1093/clinchem/hvab216.

24. Brasen CL, Christensen H, Olsen DA, Kahns S, Andersen RF, Madsen JB, Lassen A, Kierkegaard H, Jensen A, Sydenham TV, Madsen JS, Moller JK, Brandslund I. 2021. Daily monitoring of viral load measured as SARS-CoV-2 antigen and RNA in blood, IL-6, CRP and complement C3d predicts outcome in patients hospitalized with COVID-19. Clin Chem Lab Med 59:1988–1997.

25. Perna F, Bruzzaniti S, Piemonte E, Maddaloni V, Atripaldi L, Sale S, Sanduzzi A, Nicastro C, Pepe N, Bifulco M, Matarese G, Galgani M, Atripaldi L. 2021. Serum levels of SARS-CoV-2 nucleocapsid antigen associate with inflammatory status and disease severity in COVID-19 patients. Clin Immunol 226:108720.

26. Zhang Y, Ong CM, Yun C, Mo W, Whitman JD, Lynch KL, Wu AHB. 2021. Diagnostic Value of Nucleocapsid Protein in Blood for SARS-CoV-2 Infection. Clin Chem doi:10.1093/clinchem/hvab148.

27. Olea B, Albert E, Torres I, Gozalbo-Rovira R, Carbonell N, Ferreres J, Poujois S, Costa R, Colomina J, Rodriguez-Diaz J, Blasco ML, Navarro D. 2021. SARS-CoV-2 N-antigenemia in critically ill adult COVID-19 patients: Frequency and association with inflammatory and tissue-damage biomarkers. J Med Virol doi:10.1002/jmv.27300.

28. Thudium RF, Stoico MP, Hogdall E, Hogh J, Krarup HB, Larsen MAH, Madsen PH, Nielsen SD, Ostrowski SR, Palombini A, Rasmussen DB, Foged NT. 2021. Early Laboratory Diagnosis of COVID-19 by Antigen Detection in Blood Samples of the SARS-CoV-2 Nucleocapsid Protein. J Clin Microbiol 59:e0100121.

29. Li T, Wang L, Wang H, Li X, Zhang S, Xu Y, Wei W. 2020. Serum SARS-COV-2 Nucleocapsid Protein: A Sensitivity and Specificity Early Diagnostic Marker for SARS-COV-2 Infection. Front Cell Infect Microbiol 10:470.

30. Lippi G, Simundic AM, Plebani M. 2020. Potential preanalytical and analytical vulnerabilities in the laboratory diagnosis of coronavirus disease 2019 (COVID-19). Clin Chem Lab Med 58:1070–1076.

31. Basile K, McPhie K, Carter I, Alderson S, Rahman H, Donovan L, Kumar S, Tran T, Ko D, Sivaruban T, Ngo C, Toi C, O’Sullivan MV, Sintchenko V, Chen SCA, Maddocks S, Dwyer DE, Kok J. 2020. Cell-based culture of SARS-CoV-2 informs infectivity and safe de-isolation assessments during COVID-19. medRxiv doi:10.1101/2020.07.14.20153981:2020.07.14.20153981.

32. Jaafar R, Aherfi S, Wurtz N, Grimaldier C, Van Hoang T, Colson P, Raoult D, La Scola B. 2021. Correlation Between 3790 Quantitative Polymerase Chain Reaction-Positives Samples and Positive Cell Cultures, Including 1941 Severe Acute Respiratory Syndrome Coronavirus 2 Isolates. Clin Infect Dis 72:e921.

33. Favresse J, Eucher C, Elsen M, Tre-Hardy M, Dogne JM, Douxfils J. 2020. Clinical Performance of the Elecsys Electrochemiluminescent Immunoassay for the Detection of SARS-CoV-2 Total Antibodies. Clin Chem 66:1104–1106.

